# SGLT2 inhibition in addition to lifestyle intervention and risk for complications in subtypes of patients with prediabetes - a randomized, placebo controlled, multi-center trial (LIFETIME) – rationale, methodology and design

**DOI:** 10.1101/2023.11.18.23298622

**Authors:** Reiner Jumpertz von Schwartzenberg, Norbert Stefan, Robert Wagner, Martina Guthoff, Arvid Sandforth, Andrea Icks, Matthias Blüher, Jochen Seissler, Julia Szendrödi, Michael Roden, Christoph Wanner, Hiddo J. L. Heerspink, Hubert Preissl, Andreas Fritsche, Andreas L. Birkenfeld

**Affiliations:** Institute for Diabetes Research and Metabolic Diseases of the Helmholtz Center Munich at the University of Tübingen, Tübingen, Germany; German Center for Diabetes Research (DZD), Neuherberg, Germany; Department of Internal Medicine, Division of Endocrinology, Diabetology and Nephrology, University Clinic Tübingen, Tübingen, Germany; Division of Endocrinology and Diabetology, Medical Faculty, Heinrich-Heine University, Düsseldorf, Germany; Institute for Clinical Diabetology, German Diabetes Center, Leibniz Center for Diabetes Research at Heinrich-Heine University, Düsseldorf, Germany; Institute for Health Services Research and Health Economics, Centre for Health and Society, German Diabetes Center, Leibniz Center for Diabetes Research, Heinrich Heine University Düsseldorf, Düsseldorf, Germany; Institute for Health Services Research and Health Economics, German Diabetes Center, Leibniz Center for Diabetes Research, Heinrich Heine University Düsseldorf, Düsseldorf, Germany; Helmholtz Institute for Metabolic, Obesity and Vascular Research (HI-MAG) of the Helmholtz Zentrum München at the University of Leipzig and University Hospital Leipzig, Leipzig, Germany; Diabetes Research Group, Medical Department 4, Ludwig-Maximilians University Munich, Munich, Germany; Department of Endocrinology and Clinical Chemistry, Heidelberg University Hospital, Germany; Division of Nephrology, University Hospital of Würzburg, Würzburg, Germany; The George Institute for Global Health, University of New South Wales, Sydney, NSW, Australia; Department of Clinical Pharmacy and Pharmacology, University of Groningen, University Medical Center, Groningen, Netherlands

**Keywords:** SGLT2 Inhibitor, dapagliflozin, type 2 diabetes, chronic kidney disease, Tübingen Prediabetes Clusters, moderately increased albuminuria

## Abstract

**Introduction:** Type 2 diabetes (T2D) is associated with severe complications, including chronic kidney disease (CKD), cardiovascular disease, heart failure and premature death. By the time T2D is diagnosed, many patients already have emerging complications. Therefore, early treatment has the potential for prevention or even early reversal of serious complications. However, until recently, it was not well-defined which patients were most at risk of developing complications in the stage of prediabetes. By establishing the Tübingen Prediabetes Clusters, we have categorized patients with prediabetes according to their geno- and phenotype and predicted their risk for T2D and future complications. To date, no effective drug therapy has been tested in the Tübingen Prediabetes Clusters, which are at high risk of developing complications. Sodium-dependent glucose co-transporter-2 inhibitors (SGLT2) are effective in lowering blood glucose and preventing renal and cardiovascular complications in patients with T2D, heart failure or chronic higher stage renal disease. Therefore, the aim of this study is to investigate whether SGLT2 inhibition with dapagliflozin compared to placebo is effective in reducing the risk of CKD progression and other complications in patients with prediabetes from the high-risk Tübingen Prediabetes Clusters.

**Methods:** LIFETIME (NCT06054035) is a randomized, placebo-controlled, double-blind phase IV trial of 24 months duration in 182 adults within the Tübingen Prediabetes Clusters with high risk for developing complications, with prediabetes according to ADA criteria and an urinary albumin ratio (uACR) of ≥ 30mg/g. Main exclusion criteria include T2D and treatment with any glucose lowering medication.

**Results:** The primary endpoint is the mean of baseline-adjusted uACR over 24 months. Secondary outcomes include the resolution of microalbuminuria (uACR < 30mg/g), as well as changes in measured and estimated glomerular filtration rate (mGFR and eGFR). Other outcomes include the interaction between Tübingen Prediabetes Clusters and intervention, remission of prediabetes and prevention of T2D, changes in body fat distribution, small vessel integrity, myocardial function, the composition of the gut microbiome, quality of life, markers for glucose regulation as well as costs and incremental cost-effectiveness ratios, and changes of risk and time preferences. With these measures, we aim to establish the feasibility and efficacy as well as health economic aspects of an early precision treatment for patients from Tübingen Prediabetes Clusters at high risk for development of complications.

**Ethics and dissemination:** The study protocol has been reviewed and approved by all local ethics committees. The results will be disseminated through conference presentations and peer-reviewed publications.

**Registration:** The LIFETIME study was registered at clinicaltrials.gov (Identifier: NCT06054035) and EudraCT (EudraCT: 2021-005721-25).

**Strengths and limitations of this study:**

- The study is the first to test the feasibility and efficacy of an early precision treatment of complications in patients in Tübingen Prediabetes Clusters with high risk for the development of complications (high-risk Tübingen Prediabetes Clusters) and moderately increased albuminuria.
- The study design allows to explore the efficacy of SGLT2 inhibition on the interaction of treatment between the different high-risk Tübingen Prediabetes Clusters with moderately increased albuminuria.
- The two-year drug administration allows to determine the rate of decline in measured GFR in patients of these high-risk Tübingen Prediabetes Clusters to estimate needed patient numbers in future cardiorenal outcome trials in a precision treatment approach.
- The study uses highly validated and broadly accepted bridging biomarker of kidney damage and function, namely the uACR and slope of measured and estimated GFR.

## Introduction

### Background and Rational

Worldwide more than 400 million people live with T2D^1^ leading to various complications, including CKD and a twofold increased mortality rate compared to patients without diabetes, and more than 50% of patients with T2D develop micro- and/or macrovascular complications during the course of the disease.^2, 3^ However, prediabetes is much more prevalent than T2D with approx. 38% of the U.S. adult population affected^4^ and these individuals develop similar complications even before the onset of T2D, which also include CKD.^5^ However, prediabetes and early stages of CKD are often undiagnosed and affected patients later present with more advanced stages of CKD.

Using clustering analysis, we have identified 6 risk clusters of diabetes and complications development, the Tübingen Prediabetes Clusters, ^5^ of which 3 are at high risk to develop complications (i.e. clusters 3, 5, 6), partly even before the onset of T2D. While clusters 3 and 5 have the highest risk to develop T2D, CKD was the most common complication among the 3 high risk clusters,^5^ opening a window of opportunity for early tailored therapeutic strategies in these patients. We will refer to Clusters 3, 5 and 6 as high-risk Tübingen Prediabetes Clusters in the rest of the paper. So far, pharmacological therapy is not indicated for patients at risk for diabetes for whom lifestyle intervention is the only therapeutic option.

SGLT2 inhibitors reduce progression of diabetic kidney disease and ischemic heart disease in patients with T2D at high and very high cardiovascular risk, in patients with heart failure with reduced and preserved ejection fraction or in patients with advanced CKD.^6-12^ Mechanisms include hemodynamic, metabolic and vascular effects. Yet, the effect on complications in patients from high-risk Tübingen Prediabetes Clusters treated with SGLT2 inhibitors have not been studied before. Moreover, SGLT2 inhibitors have not been tested for their efficacy in patients without diabetes, normal kidney function and only moderately increased albuminuria in a dedicated progressive randomized clinical trial. Pooled prespecified analysis from exploratory endpoints showed that dapagliflozin, an SGLT2 inhibitor, reduced albuminuria in patients with and without diabetes.^10^ However, no prospective data are available in patients without diabetes in a low range of albuminuria with normal or near normal GFR (Kidney Disease: Improving Global Outcomes (KDIGO) G1A2 and G2A2). A post hoc analysis from Declare-Timi58 in patients with T2D showed that dapagliflozin did not reduce uACR in patients with albuminuria in the low normal range (uACR < 10mg/g),^13^ yet, it reduced albuminuria by 20-40% when baseline uACR exceeded 30mg/g.^13, 14^ In these patients with T2D, albuminuria was reduced rapidly within 6 weeks^13, 14^, plateaued thereafter and was significantly lower after 2 years.^13, 15^ An early intervention in patients, who are at high risk for T2D based on a cluster analysis and have early signs of CKD, holds the potential to protect patients for a longer period of time against severe organ damage or to even reverse the course of the disease.

Based on data in patients with T2D, we hypothesize that dapagliflozin reduces albumin excretion in high-risk Tübingen Prediabetes Cluster patients by at least 20% compared to placebo within a treatment period of 2 years.^15-20^ Reducing uACR during 2 years has been shown to provide clinically meaningful benefit in terms of slowing progression of disease and cardiovascular morbidity and mortality.^16^

Since there is a lack of prospective studies with SGLT2 inhibitors in patients at risk for T2D with early kidney disease (KDIGO G1A2/G2A2), we will establish the efficacy of SGLT2 inhibition in this high-risk population and determine to which extend SGLT2 inhibition can reduce albuminuria (primary endpoint), improve or preserve kidney function and promote resolution of microalbuminuria (uACR < 30mg/g) (secondary endpoints). We will also determine how SGLT2 inhibition reduces the risk of metabolic and cardiovascular parameters.

### Objectives

As a <u>primary objective</u> we will test if albuminuria in patients with prediabetes who belong to high-risk Tübingen Prediabetes Clusters 3, 5, or 6 and CKD stage G1A2/G2A2 can be reduced by a treatment with the SGLT2 inhibitor dapagliflozin (10mg/day) and lifestyle counselling compared to placebo and lifestyle counselling for two years in a double-blinded fashion.

<u>Secondary objectives</u> are to test the efficacy of the SGLT2 inhibitor dapagliflozin on kidney function and resolution of microalbuminuria (uACR < 30mg/g)

<u>Exploratory objectives</u> will test the efficacy of the SGLT2 inhibitor dapagliflozin on the resolution of prediabetes, improvement of insulin sensitivity and insulin secretion, progression to T2D as defined by HbA1c ≥ 6.5, body composition, blood pressure, onset of neuropathy, quality of life, costs and cost-effectiveness, changes of patients’ risk and time preferences. Additionally, we assess the interaction of treatment and risk cluster assignment on primary, secondary and exploratory outcomes. Further objectives are small vessel integrity assessed via vessel density and junction-to-endpoint branches, new onset or progress of retinopathy, composition of the gut microbiome, urinary metabolites and finally left ventricular mass index and systolic/diastolic myocardial function.

### Trial Design

The present study is designed as a multi-center, prospective, randomized, placebo-controlled, double-blinded, parallel group study. Patients with prediabetes from Tübingen Prediabetes Clusters 3, 5 or 6 and an early stage of CKD (KDIGO G1A2/G2A2) will be randomized to either receive lifestyle counseling plus dapagliflozin (10 mg/day) or lifestyle intervention plus placebo for a period of two years.

## Methods: Participants, Interventions, Outcome

### Study setting

The study is a multi-center trial and will be initiated in four University hospitals in Germany: Tübingen, Düsseldorf, Heidelberg and Leipzig with the lead in Tübingen. In order to meet recruitment goals, additional study sites from the German Center for Diabetes Research and other locations may be included during the course of the study.

### Eligibility Criteria

The study will include adult male and female patients aged 35 to 75 years with prediabetes and cluster assignment to the Tübingen Prediabetes Clusters 3, 5 or 6 and an early stage of CKD (KDIGO stage G1A2/G2A2). CKD stage G1A2/G2A2 include patients with a GFR ≥60 ml/min/1.73m^2^ and uACR of 30-300 mg/g. Prediabetes is defined by a fasting glucose of ≥100 mg/dl and/or 2 h glucose during a 75g oral glucose tolerance test (oGTT) ≥140 mg/dl and/or a HbA1c ≥5.7% but not meeting criteria for T2D according to current ADA guidelines.^21^ Further inclusion criteria are a body mass index ≥20 kg/m^2^, TSH within normal range, the ability to understand and follow study-related instructions and a negative pregnancy test for premenopausal women. Patients who are receiving thyroid replacement therapy must be on a stable treatment (i.e. TSH within normal range) regimen for at least 3 months prior to the screening visit. Patients who are receiving antihypertensive medication such as mineralocorticoid receptor antagonists must be on a stable treatment regimen (i.e. no hospital admission due to excessive hypertension and stable dosing) for at least 6 weeks prior to the screening visit. Patients who are treated with antihypertensive medication such as ACE inhibitors and AT1 receptor antagonists, thiazides as well as loop diuretics must be on stable treatment (i.e. no hospital admission due to excessive hypertension and stable dosing) for at least 2 weeks. Each patient has to understand and voluntarily sign an informed consent document prior to any study related assessments/procedures. The trial population will consist of both genders. No gender ratio has been stipulated in this trial as the results of clinical studies did not indicate any gender-specific differences in the effect of SGLT2 inhibitors on renal endpoints in patients with CKD.^6^

Volunteers will be excluded in case of manifest type 1 and type 2 diabetes, eGFR <60 ml/min/1.73 m^2^, intake of glucose altering medications (including current therapy with dapagliflozin or empagliflozin or any other SGLT2 inhibitor), previous therapy with dapagliflozin or other drugs that can potentially lead to overlapping toxicities within five times the half-life of the drug, known or suspected orthostatic proteinuria, any acute severe or chronic severe illness (including the following: malignant disease ongoing or < 5 years ago, unstable cardiovascular disease or revascularization procedure within 3 months prior to enrolment or expected to require coronary revascularisation procedure, acute pancreatic disease (i.e. elevated lipase 3x ULN), presence of history of or therapy for severe congestive heart failure (NYHA III and IV), pacemaker or aortic stenosis > II°, rapidly progressing or underlying kidney disease or anuria, known HIV infection or positive HIV test at screening, history of or planned organ transplantation, a history or presence of inflammatory bowel disease or other severe gastrointestinal diseases (particularly those which may impact gastric emptying, such as gastroparesis or pyloric stenosis), relevant hepatic disease, treatment with glucocorticoids, antibiotic treatment within the last 4 weeks, a history of ketoacidosis, repeated urogenital infection or acute urogenital infection at screening, hemoglobinopathies, haemolytic anaemia, or chronic anaemia (haemoglobin concentration < 12.0 g/dL), presence of psychiatric disorders or intake of antidepressant or antipsychotic agents, positive screening for severe depression (BDI ≥29), history of hypersensitivity to the study drug or its ingredients, allergy to iodine contrast dye, > 5% weight loss in the last 3 months, pregnant or breastfeeding women or any other clinical condition that would jeopardize participants’ safety or well-being while participating in this clinical trial. Furthermore, participants will have to use highly effective contraceptive methods during treatment and for 14 days (male or female) after the end of treatment and have to agree to be informed about accidental findings. Furthermore, patients will not be included in case of a current participation in other interventional clinical trial or previous treatment with other IMPs within five times the half-life of the drug.

### Intervention

The intervention consists of the SGLT2 inhibitor dapagliflozin (Forxiga®, 10mg oral once daily for 2 years) or matching placebo in addition to lifestyle intervention. The choice of the SGLT2 inhibitor and dose is based on a previous large intervention study showing benefits in patients with CKD.^6^ We chose a two year intervention period since a reduction in uACR over this period has been shown to induce a clinically meaningful benefit regarding progression of kidney disease,^15^ cardiovascular morbidity and mortality^16^ and dapagliflozin has shown beneficial effects on uACR over this period even in the lower uACR range.^13^ The control intervention consists of a placebo medication combined with lifestyle intervention. The control condition is in line with current guidelines and will therefore also benefit patients participating in the trial. In both groups, patients will undergo a conventional lifestyle intervention consisting of an in depth dietary counselling session and counselling to increase moderate-intensity physical activity as recommended according to current clinical guidelines for the prevention or delay of T2D development.^22^

### Outcomes

Primary, secondary and exploratory outcome variables are described in detail in **Table 1**.

**Table 1:**
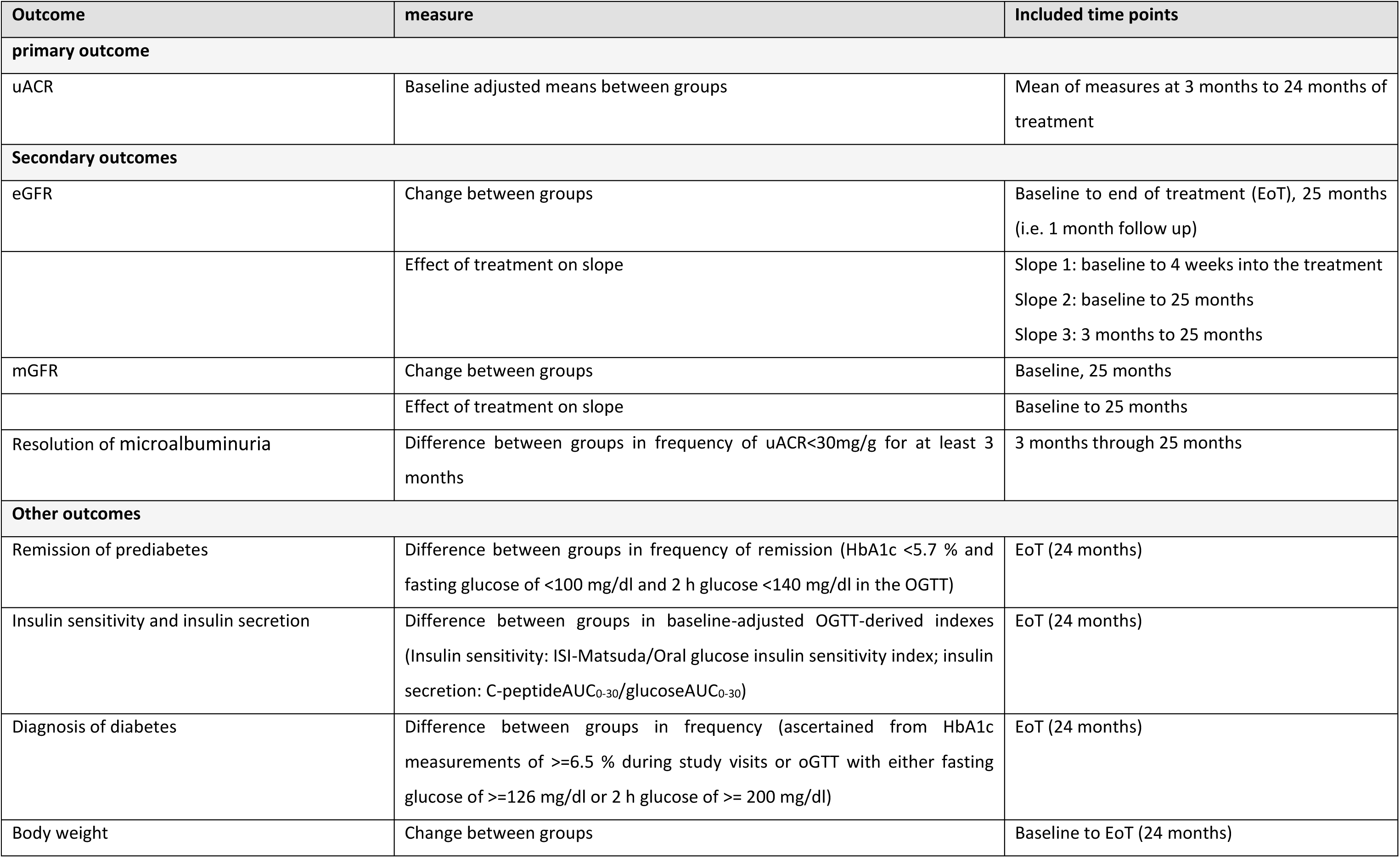
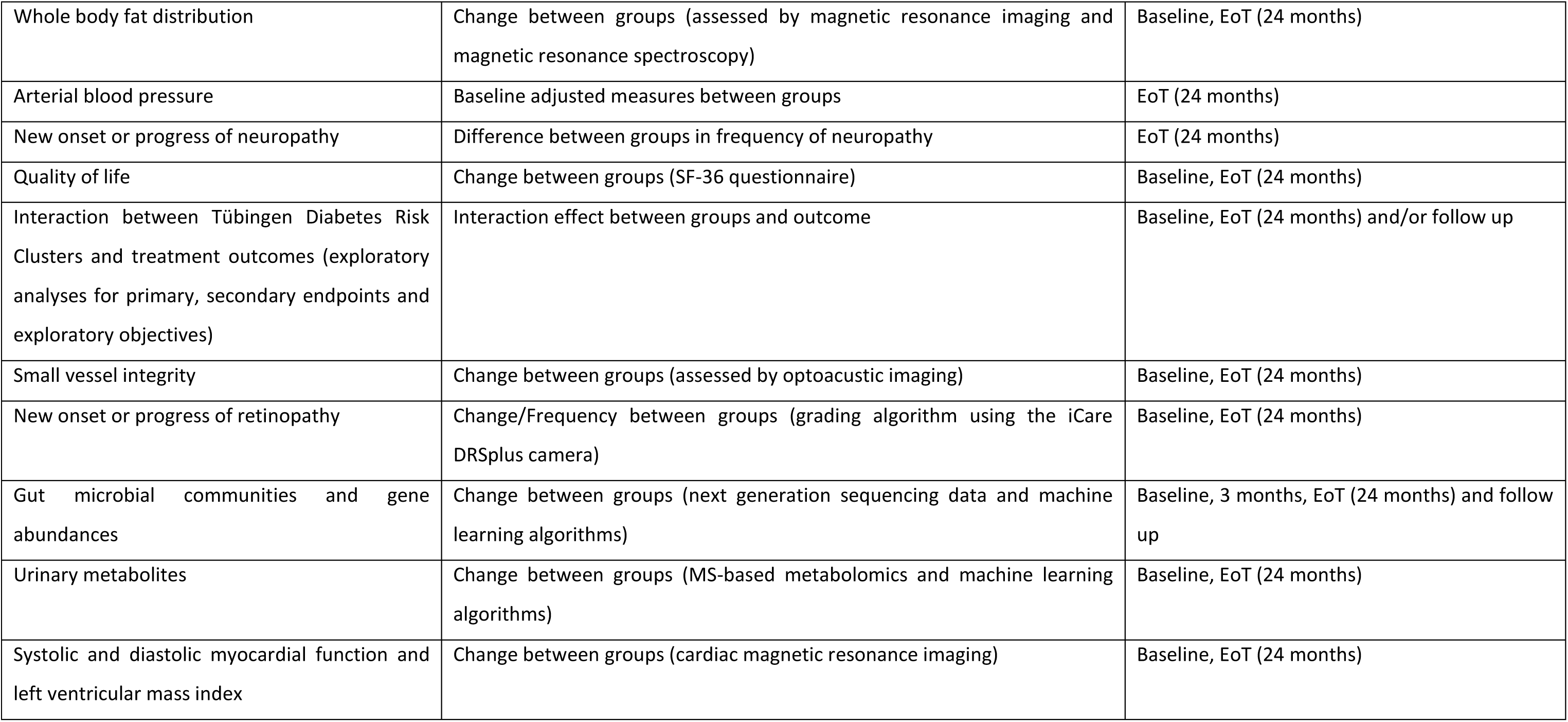
Description of outcome variables.

| Outcome | measure | Included time points |
| --- | --- | --- |
| **primary outcome** |  |  |
| uACR | Baseline adjusted means between groups | Mean of measures at 3 months to 24 months of treatment |
| **Secondary outcomes** |  |  |
| eGFR | Change between groups | Baseline to end of treatment (EoT), 25 months (i.e. 1 month follow up) |
|  | Effect of treatment on slope | Slope 1: baseline to 4 weeks into the treatment<br>Slope 2: baseline to 25 months<br>Slope 3: 3 months to 25 months |
| mGFR | Change between groups | Baseline, 25 months |
|  | Effect of treatment on slope | Baseline to 25 months |
| Resolution of microalbuminuria | Difference between groups in frequency of uACR<30mg/g for at least 3 months | 3 months through 25 months |
| **Other outcomes** |  |  |
| Remission of prediabetes | Difference between groups in frequency of remission (HbA1c <5.7 % and fasting glucose of <100 mg/dl and 2 h glucose <140 mg/dl in the OGTT) | EoT (24 months) |
| Insulin sensitivity and insulin secretion | Difference between groups in baseline-adjusted OGTT-derived indexes (Insulin sensitivity: ISI-Matsuda/Oral glucose insulin sensitivity index; insulin secretion: C-peptideAUC<sub>0-30</sub>/glucoseAUC<sub>0-30</sub>) | EoT (24 months) |
| Diagnosis of diabetes | Difference between groups in frequency (ascertained from HbA1c measurements of ≥6.5 % during study visits or oGTT with either fasting glucose of ≥126 mg/dl or 2 h glucose of ≥ 200 mg/dl) | EoT (24 months) |
| Body weight | Change between groups | Baseline to EoT (24 months) |
| Whole body fat distribution | Change between groups (assessed by magnetic resonance imaging and magnetic resonance spectroscopy) | Baseline, EoT (24 months) |
| Arterial blood pressure | Baseline adjusted measures between groups | EoT (24 months) |
| New onset or progress of neuropathy | Difference between groups in frequency of neuropathy | EoT (24 months) |
| Quality of life | Change between groups (SF-36 questionnaire) | Baseline, EoT (24 months) |
| Interaction between Tübingen Diabetes Risk Clusters and treatment outcomes (exploratory analyses for primary, secondary endpoints and exploratory objectives) | Interaction effect between groups and outcome | Baseline, EoT (24 months) and/or follow up |
| Small vessel integrity | Change between groups (assessed by optoacoustic imaging) | Baseline, EoT (24 months) |
| New onset or progress of retinopathy | Change/Frequency between groups (grading algorithm using the iCare DRSplus camera) | Baseline, EoT (24 months) |
| Gut microbial communities and gene abundances | Change between groups (next generation sequencing data and machine learning algorithms) | Baseline, 3 months, EoT (24 months) and follow up |
| Urinary metabolites | Change between groups (MS-based metabolomics and machine learning algorithms) | Baseline, EoT (24 months) |
| Systolic and diastolic myocardial function and left ventricular mass index | Change between groups (cardiac magnetic resonance imaging) | Baseline, EoT (24 months) |

### Participant Timeline

The study consists of 9 ambulatory visits: screening visit, baseline visit, treatment visits after 1, 3, 6, 12, 18 and 24 months and one follow up visit at 25 months (i.e. approx. 4 weeks after end of treatment). An outline of the study visits is given in **Figure 1**. Assessments and examinations for each visit are depicted in **Table 2**. Harmonized standard operating procedures are established for each measurement across all study sites. The estimated primary completion date of the LIFETIME study is Sept 2026.

**Figure 1:**
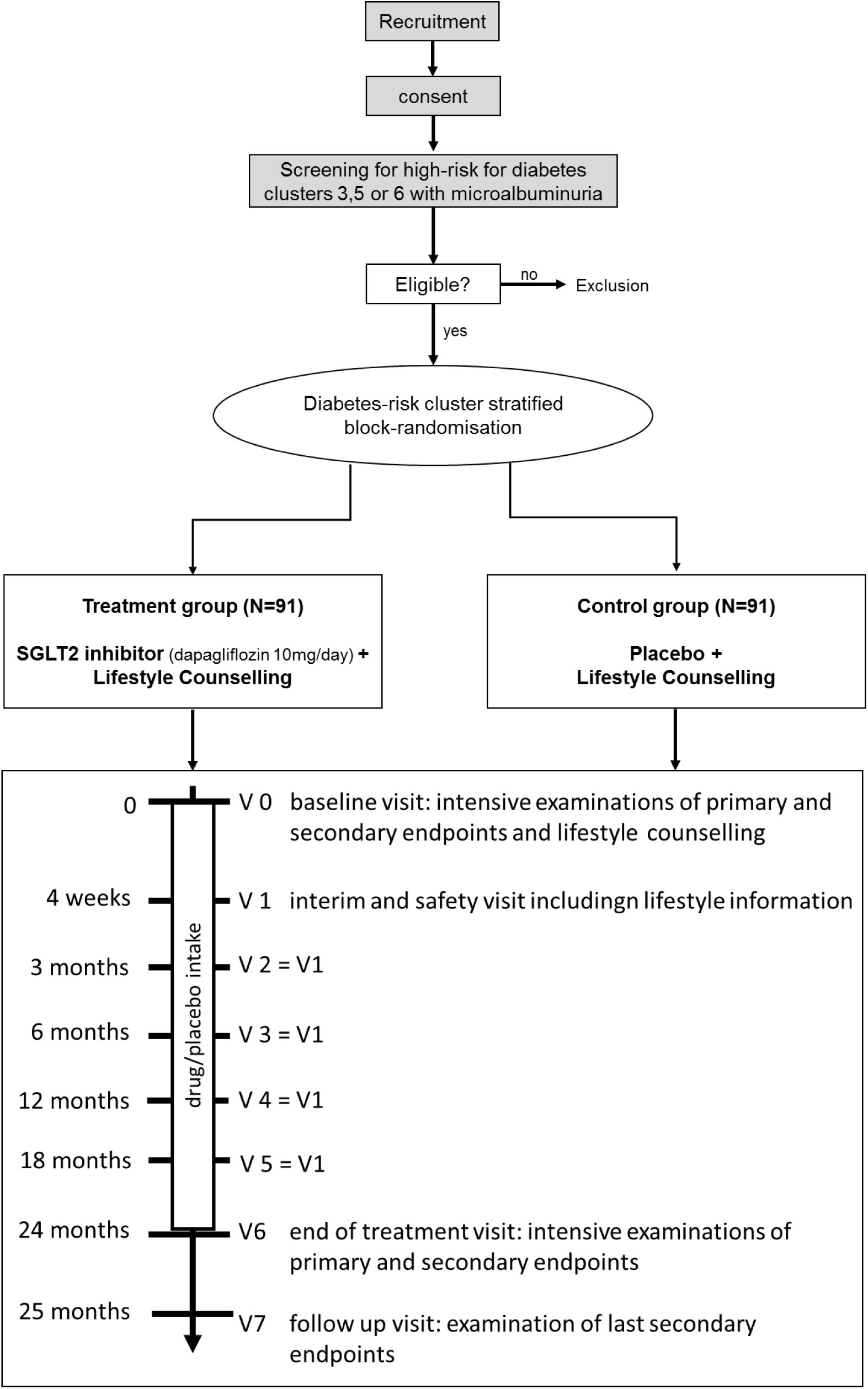
Overall Study Design.

**Table 2:** Visit schedule and study-related measures.

|  | V-1<br>Screening | V0<br>Baseline | V1 | V2 | V3 | V4 | V5 | V6<br>EoT | V7 (EoS)<br>Follow up |
| --- | --- | --- | --- | --- | --- | --- | --- | --- | --- |
| **Month** | -1 to -4 weeks<br>before baseline | 0 | 1 | 3 | 6 | 12 (1y) | 18 | 24 (2y) | 25 |
| **Written informed consent** | X |  |  |  |  |  |  |  |  |
| **Check of eligibility** | X |  |  |  |  |  |  |  |  |
| **Demographic data** | X |  |  |  |  |  |  |  |  |
| **Medical history/co-morbidities** | X |  |  |  |  |  |  |  |  |
| **Prior and concomitant medication** | X | X | X | X | X | X | X | X | X |
| **Physical examinations** | X |  |  |  |  |  |  |  |  |
| **Anthropometry and bioimpedance analysis** | X | X | X | X | X | X | X | X | X |
| **Glucose tolerance test (75g)** | X |  |  |  |  |  |  | X |  |
| **Pregnancy tests (β-HCG) in women** | X | X | X | X | X | X | X | X | X |
| **Vital signs (pulse, blood pressure)** | X | X | X | X | X | X | X | X | X |
| **Screening and Safety laboratory examinations** | X | X | X | X | X | X | X | X | X |
| **Urin analysis (assessment of uACR)** | X | X | X | X | X | X | X | X | X |
| **Screening, Safety and Clinical endpoint**<br>**laboratory examinations** |  | X |  |  |  |  |  | X |  |
| **MR-measurements** |  | X |  |  |  |  |  | X |  |
| **Optoacoustic tomography (RSOM)** |  | X |  |  |  |  |  | X |  |
| **Ocular fundus imaging (diabetic retinopathy**<br>**screening)** |  | X |  |  |  |  |  | X |  |
| **eGFR** | X | X | X | X | X | X | X | X | X |
| **mGRF** |  | X |  |  |  |  |  |  | X |
| **Rydel-Seiffer tuning fork, 10g monofilament** |  | X |  |  |  |  |  | X |  |
| **Assignment to treatment/randomization** | X |  |  |  |  |  |  |  |  |
| **Drug dispense/return/accountability** |  | X | X | X | X | X | X | X |  |
| **Lifestyle counselling/ Standard care information on a healthy lifestyle** |  | X | X | X | X | X | X |  |  |
| **Administration of study drug** |  | X |  |  |  |  |  |  |  |
| **AEs und SAEs** |  | X | X | X | X | X | X | X | X |
| **Questionnaires** | X | X |  |  |  |  |  | X |  |
| **Stool samples** |  | X |  | X |  |  |  | X | X |

### Sample Size

For the primary endpoint uACR will be analyzed in an intention-to-treat analysis. Power calculation was based on within-group variance from data of studies with a similar participant profile (derived from the previous risk cluster analysis^5^) and a corresponding uACR-range (30-299 mg/g). In a metaanalysis of the effects of SGLT2 inhibition in patients with overt diabetes, a reduction of uACR by 25-40 % was reported. ^23^ A post-hoc analysis from a trial assessing the effect of the SGLT2 inhibitor dapagliflozin in patients with severe CKD with severely increased albuminuria showed that the reduction in uACR with dapagliflozin increased with increasing HbA1c from 15% at an HbA1c of 5,7% to approx. 25% at an HbA1c of 6,4%.^10^ In our target group of participants with prediabetes, we assume a uACR reduction of 20% in the treatment group. Due to a right-skewed distribution of uACR values, the measure will be log_e_-transformed for analysis (estimated difference of means = log_e_0.8, standard deviation = 0.44). Given a power (1-β) of 80% and an ⍺ level of 0.05, we estimate an N of 64 per treatment group (pwr[package ver 1.3-0]::power.anova.test, R version 4.1.1). When we adapt our calculation by accounting for additional degrees of freedom in the ANCOVA model (baseline and center adjustment) and a drop-out rate of 30% (according to previous clinical intervention studies at our center), we aim to recruit 182 patients into the study (91 per group).

### Recruitment

Patients will be recruited from existing pools of patients and participants in previous studies, who agreed to be contacted for scientific trials as well as via email, online platforms, social media channels and advertising newspaper posts. Additionally, individuals will be recruited from the center’s outpatient clinics via the physician’s assessment of the individual risk for metabolic disease.

## Methods: Assignment of Interventions

### Allocation

Patients eligible for study participation will be randomized to one of the two treatment groups. Randomization numbers will be allocated between screening (V-1) and Visit 0 given that all results of the screening assessments are available and eligibility criteria fulfilled. Patients who fulfill the criteria of prediabetes and KDIGO stage G1A2/G2A2 but who cannot be allocated to high-risk Tübingen Prediabetes Clusters will be delayed screening failures and not randomized.

### Blinding

The study will be performed under double-blinded conditions, i.e. investigator and participants will be blinded to the treatment with dapagliflozin or placebo.

The packaging and labeling will be designed to maintain blinding to the site staff as well as to the participants. Thus, the study staff will remain blinded until database lock.

In compliance with applicable regulations, in the event of a suspected unexpected serious adverse reaction (SUSAR) the participant’s treatment code will be unblinded before reporting to the health authorities and the ethics committee.

## Methods: Data Collection, Management, Analyses

### Data Collection Methods

Participants will be randomized using a block-randomization scheme with individual randomization lists per center, risk-cluster and antihypertensive medication group (yes/no) to assure even treatment distribution within these groups.

Besides the allocation to one study arm all participants will undergo the same procedures as depicted in **Figure 1** and **Table 2**. A description of the individual procedures is given below.

#### Primary outcome variable

##### uACR

The uACR is calculated as urinary albumin in milligram per urinary creatinine in gram. Patients will provide urine samples from the morning urinary void. The mean of the screening uACR and baseline visit uACR will be taken as the baseline uACR value. For each time point, the mean of baseline adjusted values derived during visit V2-V6 will be used for the analyses.

#### Secondary outcome variable

##### eGFR-Measurement

eGFR will be calculated from routine laboratory parameters of blood samples according to the CKD-EPI 2009 formula.

##### mGFR-Measurement

For the mGFR measurement, 10 ml of an Iohexol solution approved for radiographic imaging (647.1 mg Iohexol per ml, 3235 mg Iohexol absolute, Omnipaque®) are given via an intravenous line. This is followed by a bolus of 10 ml of a commercially available saline solution. After 90, 120, 150, 180 and 210 minutes, blood samples are collected from the contralateral arm and centrifuged at 2500 g per minute for 10 minutes within 2 hours after collection. The centrifuged samples are then stored at 20°C until further analysis.

##### Resolution of microalbuminuria

Based on the uACR measure of the primary outcome variable, a resolution of microalbuminuria will be attributed to patients with a uACR < 30mg/g for at least 3 months consecutively.

#### Exploratory outcome variables

##### Oral glucose tolerance test

75-g oGTT will be used to assess glucose tolerance, whole-body insulin sensitivity and glucose-stimulated insulin secretion. On the evening before the test, participants should refrain from heavy physical activity. On the day of the testing, after an overnight fast, participants will receive orally 75 g of glucose over a period of not more than 5 min. Blood samples to assess glucose, insulin, C-peptide and proinsulin will be obtained from an indwelling antecubital venous catheter or needle at 0 min (immediately before glucose administration), 30, 60, 90, and 120 min after the glucose intake. From these measurements, established indices for insulin sensitivity and glucose-stimulated insulin secretion will be calculated. Furthermore, samples will be stored for further analyses.

##### Laboratory measurements

All laboratory values are determined in the laboratories of the respective sites except for iohexol as well as C-peptide, insulin and proinsulin measurements during OGTT that will be measured in the laboratory of the University Hospital Tübingen by an immunoassay on ADVIA Centaur XP Immunoassay System (Siemens Healthcare Diagnostics, Erlangen, Germany). For details of the laboratory assessments see **Table 3**.

**Table 3:** Screening and clinical/laboratory endpoints.

| Measure | Parameter |
| --- | --- |
| **Hematology** | B-Haemoglobin (Hb)<br>B-Leukocyte count<br>B-Leukocyte differential count (absolute count)<br>B-Platelet count |
| **Coagulation** | Quick, Partial Thromboplastin Time (PTT), Prothrombin time and International Normalized Ratio (INR) |
| **Blood Chemistry** | Albumin, Creatinine, Urea, Uric acid, Sodium, Calcium, Magnesium, Phosphate, Chloride, Potassium, Creatine Phosphokinase (CK), hsCRP, Cystatin C, Iron*, Transferrin*, Transferrin satiation*, Ferritin, IL-6, Homocystein*, Amylase*, Lipase, 25-OH vitamin D*, NT-pro BNP, folate*, vitamin B12*, HIV test<sup>§</sup> |
| **Liver Parameters** | Total bilirubin (TBL), Alanine aminotransferase (ALT), Aspartate aminotransferase (AST), Alkaline phosphatase (ALP), Gamma-glutamyl transferase (GGT), Total protein |
| **Glycemic Parameters** | FPG, HbA1c, GAD-II-antibodies, tyrosinephosphatase auto antibody (IA2 AB) |
| **Endocrine parameters** | Thyroid stimulating hormone (TSH), fT3 and fT4, Testosterone, Cortisol*, Ghrelin*, Prolactin*, beta-HCG=pregnancy test (females only) |
| **Lipid Profile** | Triglycerides (TG), Total cholesterol, HDL- and LDL-cholesterol, Free fatty acids, Lipoprotein A |
| **Urin analysis** | Dipstick for ketones, Dipstick for leukocytes and nitrite; Creatinine concentration, Urea concentration, Albumin concentration, Protein concentration |
| **OGTT <sup>#</sup>** | Blood glucose, Insulin, Proinsulin, C-peptide, Glucagon, Free fatty acids, Triglycerides |
| **Venous blood gas analysis** | pH, pO<sub>2</sub>, pCO<sub>2</sub>, base excess |

\*not at Screening; <sup>#</sup> not at V0; <sup>§</sup>only at screening

##### Anthropometric measures

Beside demographic assessments such as age, sex and ethnicity, anthropometric measures include body weight, height, and waist/hip circumference. Body mass index will be calculated by body weight (kg) / height (m^2^). Percentage of body fat and lean body mass will be determined using a Bioelectrical Impedance Analysis device (BIA) at baseline and at each follow up visit.

##### MR-based assessment of body fat distribution, ectopic lipid accumulation and cardiac function

MR examinations will be performed in the early morning after overnight fast. MR-compatible electrodes will be placed on the chest in order to derivate the ECG during cardiac MRI.^24^ The measurement consists of several short sessions starting with a few seconds localizer imaging (fast-view) for positioning of the subsequent measurements. First a localizer in three spatial orientation as well as images in 2-chamber view, 4-chamber view and short axis will be recorded. Afterwards ECG-triggered multi-slice cine imaging in short axis and single-slice cine imaging in 4-chamber view will be assessed. Mapping techniques for the quantification of T1 and T2 relaxation times will be applied.

The assessment of whole-body adipose tissue distribution consists of seamless recording of 6-7 blocks from the entire body applying a 3D chemical-shift selective imaging technique.^25^ Each block is recorded during breath-hold in 16 seconds. From this, fat- and water-selective images are calculated for quantification of adipose tissue compartments and lean tissue volumes. For the assessment of ectopic fat accumulation in the body trunk, a multi-echo Dixon imaging of abdominal organs for quantification of ectopic lipids will be recorded.^26^ After that a ^1^H-MRS of the liver will be assessed for quantification of intrahepatic lipids. For renal MRI T1- and T2-weighted anatomical images as well as functional arterial spin-labeling images will be recorded.^27^

#### Further variables

##### Ocular fundus imaging

Non-mydriatic and fully automated fundus imaging is performed with iCare DRSplus (iCare Finland Oy) fundus camera.^28^ The fully automated and quick examination permits imaging through pupils as small as 2.5 mm, without need of mydriatics. Fundus and iris are displayed by the camera and retinopathy and macula edema can be assessed from the fundus images by Artificial Intelligence eye screening software EyeArt (Eyenuk Inc., Ca, USA).^29^ Optical Coherence Tomography Angiography (OCT angiography) is carried out to assess the different layers of the retina and the retinal thickness as well as a differentiated vascular representation. This imaging examination is also non-invasive and without contrast agents.^30^

##### Neuropathy

This examination will be performed at baseline and EoT and also comprises a neuropathy screening examination using the Rydel-Seiffer tuning fork and a 10 g monofilament.

##### Raster scan optoacustic mesoscopy (RSOM)

The RSOM technique represents a novel imaging technique that allows noninvasive visualization of skin microvessel morphology and tissue perfusion with high optical contrast and resolution. The technique is based on ultrafast, low-energy laser pulses that are emitted in short succession and absorbed by endogenous chromophores (e.g., oxy- and deoxyhemoglobin). Thus, anatomical structures and pathophysiological changes can be visualized in detail. This method allows for early detection of microvessel damage.^31^

##### Stool samples

At study visits V0, V2, V6 and V7, participants will be asked to provide stool samples. Participants will be equipped with stool collection kits for self-collection at home. Additionally, participants will receive cooling pads for the sample transport to the study center. Upon arrival at the study center, samples will be aliquoted and stored at -80 °C. Samples will be analyzed for microbial composition, gene abundance as well as transcriptome and metabolomic profiles to test for effects of SGLT2 inhibitor treatment of the gut microbial communities.

##### Lifestyle counselling

All participants receive a conventional lifestyle intervention (one in depth individual session at V0 and standard care information on a healthy lifestyle at every visit thereafter). Patients will be asked to reduce fat intake to <30% of total energy intake, to reduce saturated fat intake to <10% of total energy intake, and increase intake of fibers to >15 g/1,000 kcal total energy intake. The dietary counseling will be based on diet protocols completed by the participants on four consecutive days. Additionally, participants will be encouraged to increase moderate-intensity physical activity as recommended according to current clinical guidelines for the prevention or delay of T2D development.^22^

##### Questionnaires

The Beck Depression Inventory (BDI) II assesses symptoms of depression based on the Diagnostic and Statistical Manual of Mental Disorders (DSM-IV).^32^ A commonly used eating behavior questionnaire is the Three Factor Eating Questionnaire (TFEQ) with its three domains cognitive restraint, disinhibition and experienced hunger.^33^ Physical activity of the last 12 months will be assessed via the Baecke-Score.^34^ Quality of live will be measured by the SF-36 questionnaire.^35^ For the health economic evaluation, the EQ5-D 5L will be used, as well as a questionnaire to assess health care use, which is based on a validated instrument and adjusted according to the study requirements. Risk and time preferences will be assessed by using a preference survey module.

##### Screening and Safety examinations

Routine physical examinations will be performed by a study physician including vital signs, heart, lungs and abdomen. There will be 2 blood pressure measurements made at least 30 seconds apart. Blood pressure and pulse measurement will be performed in the seated position.

**Table 3** shows the laboratory for Screening and Safety and also for clinical and laboratory endpoints. In addition, a HIV test will be performed at Screening.

### Data Management

#### Case Report Form (CRF)

Electronic Case Report Forms (eCRFs) will be used. The Clinical Data Management System (CDMS) will be used for data capture, processing and storage of study data. Data entry is performed at the investigational site by clinical staff after having received training and a user manual for the electronic CRF. Queries resulting from edit checks and/or data verification procedures will be posted electronically in the eCRF. The CDMS is validated and changes are tracked via an audit trail. The correctness of entries in CRFs will be confirmed by dated signature of an authorized investigator.

#### Source Data

Source data is all information, original records of clinical findings, observations or other activities in a clinical trial necessary for the reconstruction and evaluation of the trial. Source data are contained in source documents locally and at the distributing pharmacy.

#### Data Handling

Authorized clinical staff at the investigational site will enter the data into the eCRF using an access controlled, audit-trailed, ICH/GCP compliant, validated system. Entered data will be subjected to plausibility checks directly implemented in the CRF, monitoring and medical review. Implausible or missing data will be queried. Database lock will be performed after completion of data entry, data cleaning and a final data review.

#### Storage and Archiving of Data

According to the §13 of the German GCP-Ordinance all essential trial documents (e.g. CRF) will be archived for at least 25 years after the trial termination. The investigators will archive all trial data (source data and Investigator Site File (ISF) including subject identification list and relevant correspondence) according to the Guideline ICH GCP (E6) and to local law or regulations.

### Statistical Methods

#### Analysis Primary Variable

For the primary endpoint the parameter uACR will be analyzed as intention-to-treat analysis. This will be performed with longitudinal repeated measure analyses using mixed linear models. The dependent variable of the model will be uACR (V2-V6) with baseline uACR, center, Tübingen Prediabetes Cluster and intervention group as explanatory variables.

#### Analysis Secondary Variables

Secondary endpoints will be analyzed in the intention-to-treat population, using mixed linear models with the participant as random effect (e.g. e/mGFR trajectories). Additionally, resolution of microalbuminuria (uACR < 30mg/g) will be analyzed using Х^2^ tests.

#### Analysis Exploratory Variables

The interaction between high-risk Tübingen Prediabetes Cluster assignment and intervention will also be analyzed to test if there are efficacy differences across clusters. Further exploratory endpoints will be continuous and categorical outcomes for the endpoints listed above (see **Table 1**). Continuous secondary outcomes will be tested with linear mixed models analogous to the models above. Health economic evaluation will be conducted in form of a cost-effectiveness analysis and a cost-utility analysis. No interim analyses will be done.

No interim analyses will be done.

## Methods: Monitoring

### Data Monitoring

Monitoring for this study is provided by the Center for Clinical Studies Tübingen (Zentrum für Klinische Studien Tübingen, ZKS Tübingen). The monitoring will be conducted according to ZKS Tübingen internal Standard Operating Procedures (SOPs) and a dedicated monitoring manual for the study. The monitoring timelines include, for all centers, an initiation visit, regular monitor visits during the course of the trial as well as a close out visit. Monitoring will end with the last visit after full documentation of the last patient enrolled (close out visit).

### Harms

#### Period of Observation and Documentation

For the purpose of this trial, the period of observation for collection of adverse events (AEs) extends from the time of the patient’s first administration of the IMP until the follow up visit (V7).

AEs that occur in the course of the trial regardless of the causal relationship will be monitored and followed up until the outcome is known or no more information is achievable.

#### Documentation and Reporting of Adverse Events by the Investigator

The investigator will document all AEs that occur during the observation period set in this protocol on the pages provided in the CRF. Additional instructions may be provided in the investigator file and in the CRF itself. All serious adverse events (SAEs) reports (initial and follow-up reports), will be sent, even if they are incomplete, without undue delay but not later than within 24 hours upon receipt to a representative of the Sponsor.

#### Assessment of Severity and Causality

The investigator will also provide an assessment of the severity of the event according to CTCAE criteria (Version 5.0) and causal relationship between the event and each of the investigational products or trial procedures.

#### Sponsors Assessment of the SAEs

All SAE will be subject to a second assessment by the trial Sponsor or authorized second assessors. The second assessor will fill out a ‘Second Assessment Form’ for each SAE containing, hereby evaluating the event as serious or not, the relationship between SAE and IMP, the expectedness of SAE according to the reference documents well as a statement if the benefit / risk assessment for the trial did change as a result of SAE.

#### Follow-up of Initial Report

All patients who have adverse events, whether considered associated with the use of the investigational products or not, will be monitored to determine the outcome as far as possible. The clinical course of the adverse event will be followed up according to accepted standards of medical practice even after the end of the period of observation. The sponsor will identify missing information for each SAE report and will require follow up information in regular intervals from the investigators until all queries are resolved or no further information can be reasonably expected. All responses to queries and supply of additional information by the investigator will follow the same reporting route and timelines as the initial report.

#### Suspected Unexpected Serious Adverse Reaction (SUSAR)

All SUSARs will be reported to the responsible ethics committees, to the Bundesinstitut für Arzneimittel und Medizinprodukte (BfArM) and to all participating investigators.

#### Unblinding of the Medicinal Product / Medical Device

If it is medically imperative to know which trial medication the participant is receiving, the investigator or authorized person will unblind via CDMS. The investigator or the person who breaks the blind will record the date and the reasons for unblinding in the CRF, in the participant’s medical record on the CDMS.

### Auditing

In addition to the monitoring activities, audits will be conducted by the sponsor or assigned auditors. These audits may include checking the whole course of the study, documentation, trial center, investigators and the monitor. The competent regulatory authorities may also conduct inspections.

### Ethics and dissemination

Informed written consent is obtained from every study participant and the study is conducted in accordance to the declaration of Helsinki. The study protocol has been reviewed and approved by the ethics committee of the University Hospital Tübingen (protocol numbers 061/2023AMG1) as well as from all other sites. The results will be disseminated through conference presentations and peer-reviewed publications.

### Patients and public involvement statement

In order to better take into account the needs and interests of people living with (pre-)diabetes and to enhance the quality of research and communication we already have involved citizens and patients at different stages in the research process and have recently set up a “Citizen and Patient Advisory Board”. Furthermore, two patient representatives are included in the steering committee of the study. Annual meetings for study participants are conducted to connect patients with the responsible researchers. In addition, a newsletter with information on the latest research results is sent out once a year.

## Discussion and Outlook

This multicenter study investigates the effects of SGLT-2 inhibition with dapagliflozin in patients within high-risk clusters of prediabetes and early stages of diabetic kidney disease.

SGLT2 inhibition with dapagliflozin has so far not been tested in a dedicated prospective clinical trial in patients with prediabetes, stratified according to diabetes and complications risk. This precision medicine approach aims at providing evidence for considering patients for an early treatment who are often overlooked due to the early stage of disease.

Recent advances in precision medicine used novel clustering methods to identify different clusters of patients with T2D who harbor different risks for micro- and macrovascular complications.^36, 37^ Our group has recently expanded this concept to patients at high risk for T2D who harbor different risks for T2D development and complications partially even before the onset of T2D.^5^ These advances highlight the necessity to identify individuals with increased disease risk before the onset of complications and T2D to offer individualized treatment options. Particularly, our approach does not focus primarily on prevention of T2D in patients with prediabetes, but rather on preventing or reducing progression of complications. To our knowledge, this approach has rarely been used before, partly due to the lack of precise prediction tools of complications in people with prediabetes.

With the LIFETIME trial we address this question by also investigating the efficacy of the SGLT2 inhibitor dapagliflozin on early kidney disease in patients at increased risk of T2D. Identifying these individuals is particularly important since population-based studies have shown that even in higher CKD stages (3-5) the diagnosis is known in only 48% of affected patients.^38^ Moreover, uACR measurements are still largely missing in patients without diabetes. Importantly, not only higher CKD stages but also lower stages are rising in prevalence and large parts of this development is explained by the rising prevalence of T2D.^39^ Therefore, it is of utmost importance to identify individuals which are at risk for both, T2D and progressive CKD early in the course of the disease to offer early tailored treatment options and reduce future costs. ^40^

Protection against complications, specifically CKD, improves quality of life for patients, and reduces the individual burden of disease. For example, 30% reduction in uACR over two years reduces risk of end stage renal disease by 17-22% as well as the risk of cardiovascular mortality.^16^ Socioeconomic costs for the management of diabetes account for 37 billion EUR/year in Germany.^41^ Management of CKD in diabetes increases *per capita* costs per fiscal quarter by 3352 EUR.^41^ Reducing the severity and/or reversing CKD in patients at risk for diabetes will most likely decrease the socioeconomic burden of metabolic disease for society. Therefore, this study will also be prospectively evaluated in terms of cost-effectiveness and cost-saving qualities.

Additionally, the LIFETIME trial will set the foundation for future studies addressing specific pharmacological options for the novel Tübingen Prediabetes Clusters. Such a confirmatory trial with a novel strategy for precision medicine might show which patients benefit from an early treatment and which do not.

The multicenter setting allows for recruitment of participants from different regions within Germany and is thus not limited to a specific area. Due to the early disease stage in highly selected individuals, this study does not address endpoints such as decline of eGFR by 57%, renal mortality or end-stage renal disease, but it uses a highly validated and broadly accepted bridging biomarker for kidney disease and renal outcomes e.g. used in type 1 diabetes,^42^ namely the uACR.

In conclusion, the LIFETIME study is the first trial to study deeply phenotyped patients in different risk clusters with prediabetes defined though the Tübingen Prediabetes Clusters and mildly elevated albuminuria with the hypothesis that progression of renal and other complications can be reduced or even reversed at a very early stage in patients at high risk for diabetes by inhibition of SGLT2 using dapagliflozin. This holds the potential for improvements in the treatment of patients even before the onset of T2D and sets the stage to demonstrate the advantages of an early precise categorization of patients with different risk strata, which are very often overlooked in the primary care setting.

## Data Availability Statement

The datasets generated during the study are not available publicly because they are subject to national data protection laws and restrictions imposed by the ethics committee to ensure privacy of study participants. However, they can be requested after the primary publication through an individual project agreement with the corresponding author. The request will be reviewed by the data steering committee. A data access agreement will have to be reached after request approval.

## Abbreviations

AEs: Adverse events
CDMS: Clinical Trial Data Management System
(e)CRF: (Electronic) Case Report Form
CKD: Chronic kidney disease
e/mGFR: Estimated/measured Glomerular filtration rate
EoT: End of Treatment
KDIGO: Kidney Disease: Improving Global Outcomes
MRI: Magnetic resonance imaging
MRS: Magnetic resonance spectroscopy
OGTT: Oral glucose tolerance test
SAEs: Serious adverse events
SGLT2: Sodium-dependent glucose co-transporter-2
SUSAR: Suspected Unexpected Serious Adverse Reaction
T2D: Type 2 diabetes
uACR: Urinary albumin-creatinine ratio

## Acknowledgments

We thank all patients who supported the design of the study particularly the members of the Citizen and Patient Advisory Board of the German Center for Diabetes Research (DZD e.V.) and the patient representatives who are willing to participate in the steering committee of the LIFETEME study.

## Author contributions

RJvS and ALB drafted the manuscript. All other authors contributed to concept of the study. The study was initiated by ALB and RJvS. All authors contributed discussion of the manuscript and approved the final version before submission.

## Literature

1. World Health Organization. Diabetes 2023 [Available from: https://www.who.int/health-topics/diabetes#tab=tab_1.

2. Eizirik DL, Pasquali L, Cnop M. Pancreatic beta-cells in type 1 and type 2 diabetes mellitus: different pathways to failure. Nat Rev Endocrinol 2020;16(7):349–62. doi: 10.1038/s41574-020-0355-7

3. Braunwald E. Diabetes, heart failure, and renal dysfunction: The vicious circles. Prog Cardiovasc Dis 2019;62(4):298–302. doi: 10.1016/j.pcad.2019.07.003

4. US Centers for Disease Control and Prevention. National Diabetes Statistics Report 2022 [Available from: https://www.cdc.gov/diabetes/data/statistics-report/index.html.

5. Wagner R, Heni M, Tabak AG, et al. Pathophysiology-based subphenotyping of individuals at elevated risk for type 2 diabetes. Nat Med 2021;27(1):49–57. doi: 10.1038/s41591-020-1116-9

6. Heerspink HJL, Stefansson BV, Correa-Rotter R, et al. Dapagliflozin in Patients with Chronic Kidney Disease. N Engl J Med 2020;383(15):1436–46. doi: 10.1056/NEJMoa2024816

7. Butt JH, Nicolau JC, Verma S, et al. Efficacy and safety of dapagliflozin according to aetiology in heart failure with reduced ejection fraction: insights from the DAPA-HF trial. Eur J Heart Fail 2021;23(4):601–13. doi: 10.1002/ejhf.2124 [published Online First: 20210310]

8. McMurray JJV, Solomon SD, Inzucchi SE, et al. Dapagliflozin in Patients with Heart Failure and Reduced Ejection Fraction. N Engl J Med 2019;381(21):1995–2008. doi: 10.1056/NEJMoa1911303

9. Jhund PS, Claggett BL, Talebi A, et al. Effect of Dapagliflozin on Total Heart Failure Events in Patients With Heart Failure With Mildly Reduced or Preserved Ejection Fraction: A Prespecified Analysis of the DELIVER Trial. JAMA Cardiol 2023;8(6):554–63. doi: 10.1001/jamacardio.2023.0711

10. Jongs N, Greene T, Chertow GM, et al. Effect of dapagliflozin on urinary albumin excretion in patients with chronic kidney disease with and without type 2 diabetes: a prespecified analysis from the DAPA-CKD trial. Lancet Diabetes Endocrinol 2021;9(11):755–66. doi: 10.1016/S2213-8587(21)00243-6 [published Online First: 20211004]

11. Heerspink HJL, Jongs N, Chertow GM, et al. Effect of dapagliflozin on the rate of decline in kidney function in patients with chronic kidney disease with and without type 2 diabetes: a prespecified analysis from the DAPA-CKD trial. Lancet Diabetes Endocrinol 2021;9(11):743–54. doi: 10.1016/S2213-8587(21)00242-4 [published Online First: 20211004]

12. Brown E, Heerspink HJL, Cuthbertson DJ, et al. SGLT2 inhibitors and GLP-1 receptor agonists: established and emerging indications. Lancet 2021;398(10296):262–76. doi: 10.1016/S0140-6736(21)00536-5

13. Mosenzon O, Wiviott SD, Heerspink HJL, et al. The Effect of Dapagliflozin on Albuminuria in DECLARE-TIMI 58. Diabetes Care 2021;44(8):1805–15. doi: 10.2337/dc21-0076

14. Pollock C, Stefansson B, Reyner D, et al. Albuminuria-lowering effect of dapagliflozin alone and in combination with saxagliptin and effect of dapagliflozin and saxagliptin on glycaemic control in patients with type 2 diabetes and chronic kidney disease (DELIGHT): a randomised, double-blind, placebo-controlled trial. Lancet Diabetes Endocrinol 2019;7(6):429–41. doi: 10.1016/S2213-8587(19)30086-5

15. Cherney DZI, Zinman B, Inzucchi SE, et al. Effects of empagliflozin on the urinary albumin-to-creatinine ratio in patients with type 2 diabetes and established cardiovascular disease: an exploratory analysis from the EMPA-REG OUTCOME randomised, placebo-controlled trial. The Lancet Diabetes & Endocrinology 2017;5(8):610–21. doi: 10.1016/s2213-8587(17)30182-1

16. Coresh J, Heerspink HJL, Sang Y, et al. Change in albuminuria and subsequent risk of end-stage kidney disease: an individual participant-level consortium meta-analysis of observational studies. Lancet Diabetes Endocrinol 2019;7(2):115–27. doi: 10.1016/S2213-8587(18)30313-9

17. Wanner C, Inzucchi SE, Lachin JM, et al. Empagliflozin and Progression of Kidney Disease in Type 2 Diabetes. N Engl J Med 2016;375(4):323–34. doi: 10.1056/NEJMoa1515920

18. Zinman B, Wanner C, Lachin JM, et al. Empagliflozin, Cardiovascular Outcomes, and Mortality in Type 2 Diabetes. N Engl J Med 2015;373(22):2117–28. doi: 10.1056/NEJMoa1504720

19. Perkovic V, Jardine MJ, Neal B, et al. Canagliflozin and Renal Outcomes in Type 2 Diabetes and Nephropathy. N Engl J Med 2019;380(24):2295–306. doi: 10.1056/NEJMoa1811744

20. Wiviott SD, Raz I, Bonaca MP, et al. Dapagliflozin and Cardiovascular Outcomes in Type 2 Diabetes. N Engl J Med 2019;380(4):347–57. doi: 10.1056/NEJMoa1812389

21. ElSayed NA, Aleppo G, Aroda VR, et al. 2. Classification and Diagnosis of Diabetes: Standards of Care in Diabetes-2023. Diabetes Care 2023;46(Suppl 1):S19-S40. doi: 10.2337/dc23-S002

22. VersorgungsLeitlinie N. Therapie des Typ-2-Diabetes Langfassung 1. 2013;1(4)

23. Piperidou A, Sarafidis P, Boutou A, et al. The effect of SGLT-2 inhibitors on albuminuria and proteinuria in diabetes mellitus: a systematic review and meta-analysis of randomized controlled trials. J Hypertens 2019;37(7):1334–43. doi: 10.1097/Hjh.0000000000002050

24. Thiele H, Nagel E, Paetsch I, et al. Functional cardiac MR imaging with steady-state free precession (SSFP) significantly improves endocardial border delineation without contrast agents. J Magn Reson Imaging 2001;14(4):362–7. doi: 10.1002/jmri.1195

25. Haueise T, Schick F, Stefan N, et al. Analysis of volume and topography of adipose tissue in the trunk: Results of MRI of 11,141 participants in the German National Cohort. Sci Adv 2023;9(19):eadd0433. doi: 10.1126/sciadv.add0433 [published Online First: 20230512]

26. Machann J, Hasenbalg M, Dienes J, et al. Short-Term Variability of Proton Density Fat Fraction in Pancreas and Liver Assessed by Multiecho Chemical-Shift Encoding-Based MRI at 3 T. J Magn Reson Imaging 2022;56(4):1018–26. doi: 10.1002/jmri.28084 [published Online First: 20220127]

27. Rossi C, Artunc F, Martirosian P, et al. Histogram analysis of renal arterial spin labeling perfusion data reveals differences between volunteers and patients with mild chronic kidney disease. Invest Radiol 2012;47(8):490–6. doi: 10.1097/RLI.0b013e318257063a

28. Sarao V, Veritti D, Borrelli E, et al. A comparison between a white LED confocal imaging system and a conventional flash fundus camera using chromaticity analysis. BMC Ophthalmol 2019;19(1):231. doi: 10.1186/s12886-019-1241-8 [published Online First: 20191119]

29. Ipp E, Liljenquist D, Bode B, et al. Pivotal Evaluation of an Artificial Intelligence System for Autonomous Detection of Referrable and Vision-Threatening Diabetic Retinopathy. JAMA Netw Open 2021;4(11):e2134254. doi: 10.1001/jamanetworkopen.2021.34254 [published Online First: 20211101]

30. Matsunaga DR, Yi JJ, De Koo LO, et al. Optical Coherence Tomography Angiography of Diabetic Retinopathy in Human Subjects. Ophthalmic Surg Lasers Imaging Retina 2015;46(8):796–805. doi: 10.3928/23258160-20150909-03

31. Haedicke K, Agemy L, Omar M, et al. High-resolution optoacoustic imaging of tissue responses to vascular-targeted therapies. Nat Biomed Eng 2020;4(3):286–97. doi: 10.1038/s41551-020-0527-8 [published Online First: 20200312]

32. Beck AT, Steer RA, Garbin MG. Psychometric Properties of the Beck Depression Inventory - 25 Years of Evaluation. Clin Psychol Rev 1988;8(1):77–100. doi: Doi 10.1016/0272-7358(88)90050-5

33. Pudel V, Westenhöfer, J. Fragebogen zum Eßverhalten (FEV)-Handanweisung. Göttingen: Verlag für Psychologie Dr. CJ Hogrefe 1989.

34. Baecke JA, Burema J, Frijters JE. A short questionnaire for the measurement of habitual physical activity in epidemiological studies. Am J Clin Nutr 1982;36(5):936–42. doi: 10.1093/ajcn/36.5.936

35. Ware JE, Jr., Sherbourne CD. The MOS 36-item short-form health survey (SF-36). I. Conceptual framework and item selection. Med Care 1992;30(6):473–83.

36. Zaharia OP, Strassburger K, Strom A, et al. Risk of diabetes-associated diseases in subgroups of patients with recent-onset diabetes: a 5-year follow-up study. Lancet Diabetes Endocrinol 2019;7(9):684–94. doi: 10.1016/S2213-8587(19)30187-1 [published Online First: 20190722]

37. Ahlqvist E, Storm P, Karajamaki A, et al. Novel subgroups of adult-onset diabetes and their association with outcomes: a data-driven cluster analysis of six variables. Lancet Diabetes Endocrinol 2018;6(5):361–69. doi: 10.1016/S2213-8587(18)30051-2 [published Online First: 20180305]

38. Carpio EM, Ashworth M, Asgari E, et al. Hypertension and cardiovascular risk factor management in a multi-ethnic cohort of adults with CKD: a cross sectional study in general practice. J Nephrol 2022;35(3):901–10. doi: 10.1007/s40620-021-01149-0 [published Online First: 20211116]

39. Coresh J, Selvin E, Stevens LA, et al. Prevalence of chronic kidney disease in the United States. JAMA 2007;298(17):2038–47. doi: 10.1001/jama.298.17.2038

40. Cusick MM, Tisdale RL, Chertow GM, et al. Population-Wide Screening for Chronic Kidney Disease: A Cost-Effectiveness Analysis. Ann Intern Med 2023;176(6):788–97. doi: 10.7326/M22-3228 [published Online First: 20230523]

41. Deutsche Diabetes Gesellschaft (DDG) und diabetesDE – Deutsche Diabetes-Hilfe. Deutscher Gesundheitsbericht - Diabetes 2021 - Die Bestandsaufnahme: Verlag Kirchheim + Co GmbH 2021.

42. Heerspink HJL, Birkenfeld AL, Cherney DZI, et al. Rationale and design of a randomised phase III registration trial investigating finerenone in participants with type 1 diabetes and chronic kidney disease: The FINE-ONE trial. Diabetes Res Clin Pract 2023:110908. doi: 10.1016/j.diabres.2023.110908 [published Online First: 20230914]

